# Haplotype networks of SARS-CoV-2 infections in the Diamond Princess cruise ship outbreak

**DOI:** 10.1101/2020.03.23.20041970

**Authors:** Tsuyoshi Sekizuka, Kentaro Itokawa, Tsutomu Kageyama, Shinji Saito, Ikuyo Takayama, Hideki Asanuma, Naganori Nao, Rina Tanaka, Masanori Hashino, Takuri Takahashi, Hajime Kamiya, Takuya Yamagishi, Kensaku Kakimoto, Motoi Suzuki, Hideki Hasegawa, Takaji Wakita, Makoto Kuroda

## Abstract

The Diamond Princess (DP) cruise ship was put under quarantine offshore Yokohama, Japan, after a passenger who disembarked in Hong Kong was confirmed as a COVID-19 case. We performed whole genome sequencing of SARS-CoV-2 directly from PCR-positive clinical specimens and conducted a haplotype network analysis of the outbreak. All tested isolates exhibited a transversion at G_11083_T, suggesting that SARS-CoV-2 dissemination on the DP originated from a single introduction event before the quarantine started. Although further spreading might have been prevented by quarantine, some progeny clusters were linked to transmission through mass-gathering events in the recreational areas and direct transmission among passengers who shared cabins during the quarantine. This study demonstrates the usefulness of haplotype network analysis in identifying potential infection routes.

**One Sentence Summary:** Genome-based tracing of SARS-CoV-2 infections among passengers and crews in Diamond Princess cruise ship during the quarantine

## Main Text

In late December 2019, an outbreak of a novel coronavirus disease (COVID-19) originated in Wuhan, China. It was caused by a new strain of betacoronavirus: severe acute respiratory syndrome coronavirus 2 (SARS-CoV-2) (*1, 2*). The Diamond Princess (DP) cruise ship was put under quarantine soon after its return to Yokohama, Japan on Feb. 3, 2020, after visiting Kagoshima, Hong Kong, Vietnam, Taiwan, and Okinawa, because an 80-year-old passenger who disembarked in Hong Kong was confirmed as a COVID-19 case on Feb. 1 after the ship had departed from Hong Kong on Jan. 25. The passenger presented with cough beginning on Jan. 23. From Feb. 3–4, the health status of all passengers and crew members was investigated by quarantine officers and upper-respiratory specimens were collected from symptomatic passengers, crew, and their close contacts for SARS-CoV-2 PCR testing. As of Feb. 5, there were a total of 3,711 individuals with 2,666 passengers and 1,045 crew members on board the DP. The Japanese government has asked about 3,600 passengers and crew members to stay on board in Yokohama during the 14-day isolation period through Feb. 19 to prevent the further spread of COVID-19 cases. Field briefing (DP COVID-19 Cases) has been documented (*3*). As of March 8, 697 COVID-19 cases had been identified among the 3,711 persons in total on the DP and seven people had died (*4*).

Here, we have generated a haplotype network of the SARS-CoV-2 outbreak using genome-wide single nucleotide variations (SNVs), identifying the genotypes of isolates that disseminated in the DP cruise ship after quarantine on Feb. 5. This study was approved by the research ethics committee of the National Institute of Infectious Diseases (approval no. 1091)

During the period from Feb. 15–17, we obtained the pharyngeal specimens of 896 persons (880 passengers, 15 crew members, and 1 quarantine officer). RT-qPCR testing for SARS-CoV-2 was positive for 148 individuals (138 passengers, 9 crew members, and 1 quarantine officer), indicating that the positivity rate was 16.4% (147 positive individuals [excluding the officer] / 895 subjects). This positivity rate was comparable to the rate determined after finishing PCR testing for all persons (619 PCR positives for 3,711 persons, 16,6%; see report for more detail) (*5*).

A detected Cq (cycle quantification) value of RT-qPCR for the 896 specimens is shown in Supplementary Table S1. All 148 RT-qPCR positive RNA samples were subjected to the PrimalSeq protocol to generate enriched cDNA of the SARS-CoV-2 genome. Samples were then sequenced in an Illumina NextSeq 500. In total 70 WGSs have been determined. The Cq limit for successful genome sequence determination was around 32 (Fig. 1A), which corresponds to a virus copy number of less than 300. The 73 genome sequences (including 3 sequences of DP isolates deposited in GISAID) were compared with the Wuhan-Hu-1 (isolated on Dec. 26, 2020 in China) genome sequence as a reference. The frequencies of single nucleotide variations (SNVs) suggested that all 73 isolates shared a single nucleotide variation: the G nucleotide at the 11,083 position on the Wuhan-Hu-1 genome sequence was mutated to T (G_11083_T transversion) leading to a non-synonymous amino acid substitution (Leu_37_Phe) in the nsp6 protein (Fig. 1B). Some additional minor SNVs were identified throughout the genome sequence.

**Fig 1.**
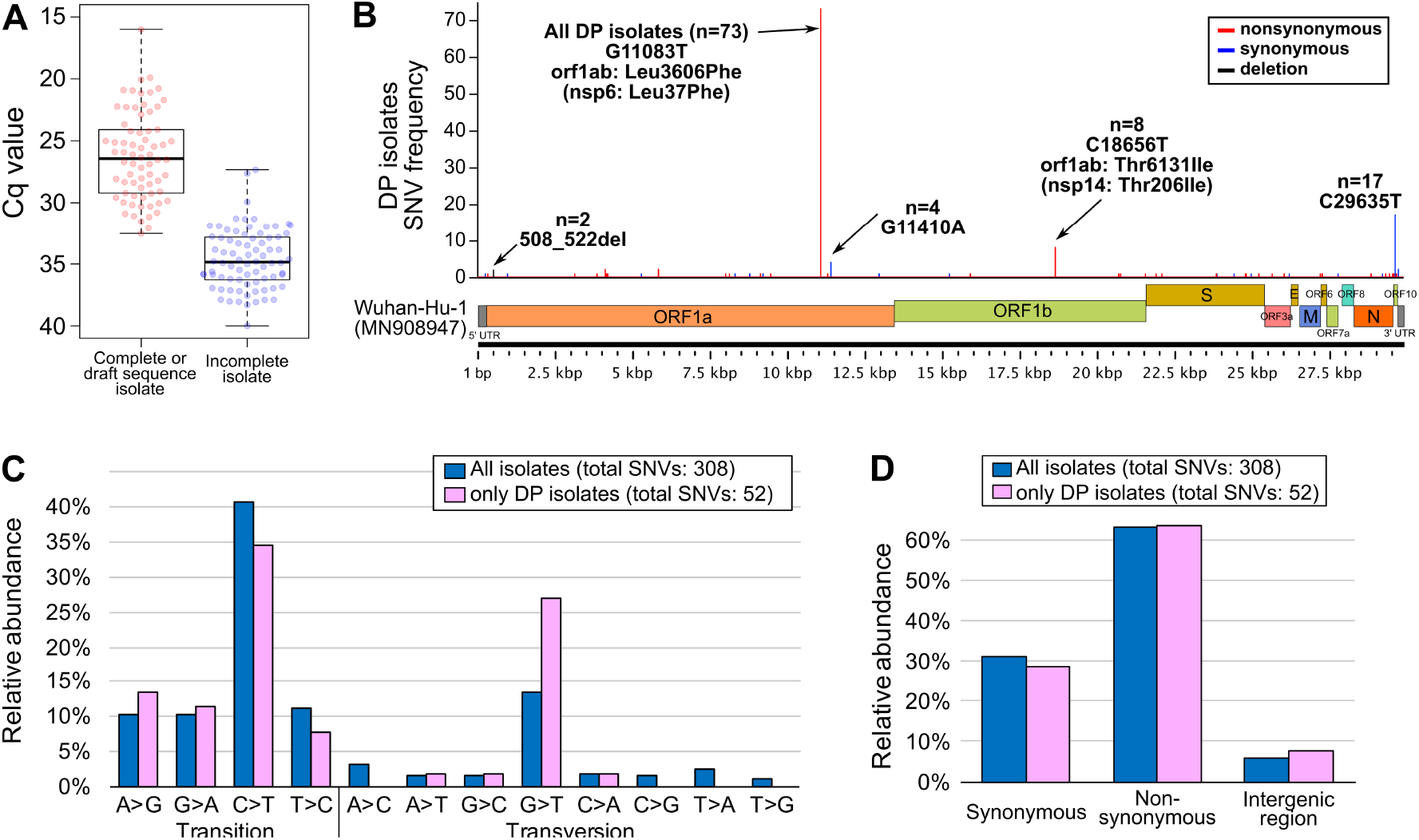
Whole genome sequence of SARS-CoV-2 isolates (strains) on the Diamond Princess (DP) cruise ship off Yokohama in Japan. A) Box plots of the Cq value from RT-qPCR assays that either successfully resulted in whole genome sequencing or incomplete sequencing. A Cq of at least 32 (∼300 viral copy) is favorable for obtaining the whole genome sequence by PrimalSeq. B) The genome position of genetic variations found among DP isolates compared with the isolate Wuhan-Hu-1 (MN908947). C) The detected nucleotide transition and transversion in all GISAID available SARS-CoV-2 genomes (n= 248, see Supplementary Table S3. Updated on March 10, 2020) and DP isolates (n=73) compared with the isolate Wuhan-Hu-1 (MN908947). D) The detected ratio of synonymous/non-synonymous mutations on coding sequences or intergenic regions in all GISAID available SARS-CoV-2 genomes (n= 248, Updated on March 10, 2020) and DP isolates (n=73) compared with the isolate Wuhan-Hu-1 (MN908947).

A number of nucleotide variations have been identified; specifically, 52 SNVs among DP isolates (n=73) and 308 SNVs among all isolates, including the Global Initiative on Sharing All Influenza Data (GISAID entries [n=248], Supplementary Table S3) (Fig. 1C). Pyrimidine transition, in particular cytosine (C) to thymine (T), makes up a large portion of the SNVs, suggesting that hydrolytic deamination of cytosine and 5-methylcytosine residues in DNA appears to contribute significantly to the appearance of spontaneous mutations (*6*). On the other hand, transversion most likely occurs at a low rate, but the guanine (G) to thymine (T) transversion in particular has been observed in both DP isolates and other SARS-CoV-2 isolates (Fig. 1C). Naturally generated reactive oxygen alters guanine to 8-oxo-guanine (8oxoG), causing significant destabilization of the 8oxoG and C base pair (*7*), implying that G to T transversion is one of possible driving forces generating SARS-CoV-2 variants. Intriguingly, the occurrence of non-synonymous mutations (63%) is apparently higher than that of synonymous mutations (31%) (Fig. 1D), suggesting that early generations of SARS-CoV-2 variants may acquire possible pivotal mutations to adapt to humans.

Haplotype networks from genomic SNVs (HN-GSNVs) were used to map the genotypes of the SARS-CoV-2 isolates that disseminated in the DP cruise ship after isolation of the passengers on Feb. 5. In addition to identifying the genetic alterations, we generated HN-GSNVs to highlight the clonality of DP isolates compared with other SARS-CoV-2 genome sequences which are publicly available on GISAID (update as of March 10, 2020). Analysis clearly suggested that the DP cluster had been generated from the potential haplotype ancestor of the global pandemic, Wuhan-Hu-1 haplotype, by G_11083_T transversion before initial dissemination in the DP (Fig. 2A, and see Supplementary Table S2 for SNVs). Thus far, all DP isolates identified have been rooted on a single brunch of the HN-GSNVs map (Fig. 2A), showing different genealogy to other isolates in China, Europe, North America, and an outbreak on another cruise ship reported from the USA. The finding that all isolates in the DP cluster exhibit G_11083_T transversion strongly suggest that SARS-CoV-2 dissemination in the DP could have originated from a single introduction event before the quarantine started on Feb. 3. Further focusing of the HN-GSNVs map on only the DP isolates in the cluster revealed that the 29 isolates of the DP-A cluster (indicated A in Fig. 2B) predominated among the DP isolates, indicating that the DP-A cluster is the ancestral haplotype for subsequent transmission. Based on the estimated speed of SARS-CoV-2 evolution (temporal resolution assumes a nucleotide substitution rate of 8 × 10^−4^ subs per site per year among the 528 SARS-CoV-2 WGSs, described at the Nextstrain web site for real-time tracking of pathogen evolution [update as of March 16, 2020] (*8*), one mutation per genome occurs in every 15 days in average, suggesting that predominant isolates in the DP-A cluster could be a very early generation of progeny from Wuhan-Hu-1 isolated on Dec. 26, 2020. This predomination of the DP-A cluster might explain the large epidemics within the cruise ship; it is possible that super-spreading occurred immediately after introduction of the virus, well before the quarantine starting on Feb. 3. This super-spreading may have originated from the COVID-19 patient who had disembarked in Hong Kong on Jan. 25. Although further explosive spreading might have been prevented after the quarantine, some of the subsequent progeny clusters, including DP-B (5 isolates) and DP-C (6 isolates) (Fig. 2B), may have formed via transmission through hidden links, such as eating at the same dinner table. Additionally, 33 patients (45%) not included in the DP-A, -B and -C clusters were revealed to have unique SARS-CoV-2 haplotypes with patient-specific unique SNVs and/or deletions. The finding of such unique progenies might explain how further subsequent minor spreading occurred after the quarantine. Indeed, the haplotype network suggests the existence of direct transmissions among passengers who shared same cabins during the quarantine; three cabin mates had been infected by the virus with same haplotype (two pairs in DP-A cluster, and one pair in other cluster), and 5 pairs shared haplotypes linked by a single SNV (A to DP0290, A to DP0481, A to DP0645, A to DP0803, and DP0764 to DP0765).

**Fig 2.**
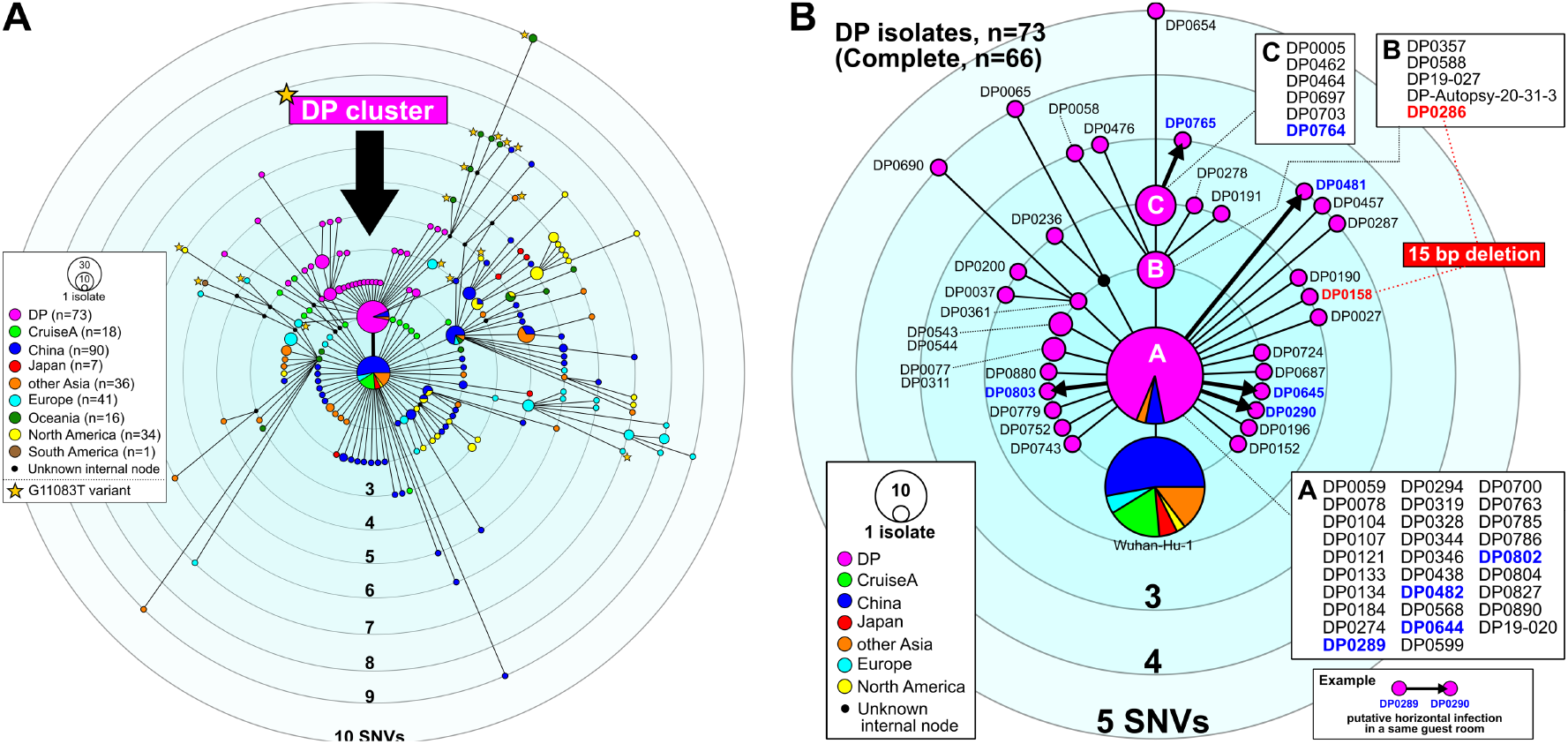
Haplotype network using genome-wide single nucleotide variations (HN-GSNVs). A) Comparative median-joining SNV network analysis obtained from whole genome sequences of 73 SARS-CoV-2 isolates (Feb. 15–17, 2020) on the Diamond Princess (DP) cruise ship off Yokohama in Japan compared with all GISAID-available SARS-CoV-2 genomes (n=248, updated on March 10, 2020). B) Clusters of the DP isolates (n=73) and Wuhan-Hu-1 were selected from the network shown in Fig. 2A to identify the strain that was first spread (indicated A, n=29 isolates) and subsequent transmissions (indicated B, n=5 isolates; indicated C, n=6 isolates; and the other 33 isolates). Information on each isolate can be obtained in Supplementary Table S1. A possible transmission between partners in same cabin after the quarantine is shown by thick arrows between isolate nodes.

We did not find any area-specific events of COVID-19 spread in the DP because all COVID-19 patients were widely distributed in all 18 decks across the ship. This may indicate that most SARS-CoV-2 infections began at mass-gathering events in the recreational areas where all passengers enjoyed dancing, singing, shopping, and watching performances. The highest incidence of COVID-19 onset was observed on February 7; the epidemic curve in the latter half of the epidemic was dominated by crew members whose work was not strictly controlled to maintain service on the ship (*9*). Nishiura suggested that the peak time of infection was from February 2–4, predicted by back-calculating the incidence of SARS-CoV-2 infections on the DP, and that the incidence abruptly declined afterwards (*9*). Nishiura also have stated that the estimated incidence indicates that the movement restriction policy was highly successful in greatly reducing the number of secondary transmissions on board the DP (*9*).

Another report by Rocklöv et al. suggested that the public health measures and quarantine prevented more than 2,000 additional cases compared to a scenario in which no interventions were taken (*10*). Rocklöv et al. did acknowledge that evacuating all passengers and crew early would have prevented many more passengers and crew from becoming infected, although this containment method was not realistic at the time. Zhang et al. also estimated that the reproduction number (R_0_) for COVID-19 cases was 2.28 (95% confidence interval: 2.06–2.52) during the early stage, and estimated that the outbreak size on the DP cruise ship might have been greater had the strict infection management and quarantine not been conducted (*11*).

An increased number of COVID-19 cases have been confirmed, and it is now difficult to identify infection routes because some Japanese cases have no recent travel history to China or contact with persons from Wuhan City or Hubei Province in China. Regarding the quarantine on the DP cruise ship, the enclosed circumstances revealed how SARS-CoV-2 can be spread in the community. Further field epidemiological studies should be conducted to trace the link of infections but, unfortunately, efforts to identify the route of transmission are still limited. This study demonstrated that HN-GSNVs can contribute to epidemiologic field investigations and identified possible transmission routes in the DP cruise ship outbreak after the quarantine was enacted.

## Data Availability

The new sequences have been deposited in GISAID with accession IDs EPI_ISL_416565 – EPI_ISL_416634. UPDATE 23/3/20

## Acknowledgments

We deeply thank the staff of the Quarantine Station for the collection and transport of clinical specimens. In particular, we are grateful to all the staff of the Influenza Virus Research Center and Department of Virology 3, National Institute of Infectious Diseases, for the laboratory testing. We thank Dr. Makoto Ohnishi and all the staff of the National Institute of Infectious Diseases, Field Epidemiology Training Program (FETP) team, the Ministry of Health, Labor and Welfare, and local governments for their assistance with administrative matters, field investigation, data collection, and laboratory testing. We would like to thank all the authors who have kindly deposited and shared genome data on GISAID. A table with genome sequence acknowledgments can be found in Supplementary Table S3. We would like to thank Editage (www.editage.com) for English language editing.

## Author Contributions

T.S., K.I., T.W. and M.K. designed and organized the genome study. R.T. and M.H. performed the genome sequencing, and T.S. and K.I. performed the genome analysis. T.K., S.S., I.T., H.A., N.N. and H.H. performed the laboratory detection. T.T., H.K., T.Y. and M.S. contributed the field epidemiological study. M.K. wrote the report.

## Data and materials availability

The new sequences have been deposited in GISAID with accession IDs EPI_ISL_416565 – EPI_ISL_416634. UPDATE 23/3/20:

## Supplementary Materials

Materials and Methods

Tables S1-S3

